# Global years of life lost to COVID-19

**DOI:** 10.1101/2020.06.19.20136069

**Authors:** Héctor Pifarré i Arolas, Enrique Acosta, Guillem López Casasnovas, Adeline Lo, Catia Nicodemo, Tim Riffe, Mikko Myrskylä

## Abstract

Understanding the mortality impact of COVID-19 requires not only counting the dead, but analyzing how premature the deaths are. We calculate years of life lost (YLL) across 42 countries due to COVID-19 attributable deaths, and also conduct an analysis based on estimated excess deaths. As of June 13th 2020, YLL in heavily affected countries are 2 to 6 times the average seasonal influenza; over two thirds of the YLL result from deaths in ages below 75 and one quarter from deaths below 55; and men have lost 47% more life years than women. The results confirm the large mortality impact of COVID-19 among the elderly. They also call for heightened awareness in devising policies that protect vulnerable demographics losing the largest number of life-years.

**One Sentence Summary:** Across 42 countries, the years of life lost due to COVID-19 are up to 6 times that of the average seasonal flu.

## Main Text

The COVID-19 pandemic is influencing all dimensions of life, from the most obvious and tragic direct health and mortality impacts to the complex individual and societal responses that are required to keep societies functioning. Policy responses, when reasonable, are a balancing act between minimizing the immediate health impact of the pandemic, and containing the long-term damage to the society that may arise from the protective policies. A key input parameter in the calculation of how restrictive policies might be justified is the mortality impact of COVID-19.

Attempts to evaluate the total mortality impact of COVID-19 are proceeding on several fronts. Progress is being made in estimating the infection fatality rate of COVID-19 and how this might vary across sub-populations *(1)*. Large, coordinated international collaborations have been set up to collect data that records COVID-19 attributable deaths. Attempts to estimate total excess mortality related to the COVID-19 are underway, and emphasized as an important measure *(2,3)*. Each of these research avenues is important in informing the public and policymakers about the mortality impact of COVID-19. However, each come with their own limitations. Infection fatality rates apply only to the relatively small sub-population that has been confirmed to have the disease, and without knowledge about the true number of infected, these rates are inherently difficult to estimate. COVID-19 attributable deaths may over- or underestimate the true number of deaths that are due to the disease, as both policies and practices about coding the deaths are only being developed and standardized. Excess death approaches that compare mortality rates during the COVID-19 outbreak to a baseline depend on correctly estimating the baseline.

The most important limitation in COVID-19 attributable death or excess death approaches, however, is that these approaches do not provide information on how many life years have been lost. Deaths at very old ages can be considered to result in fewer life years lost, when compared to deaths at very young ages. In fact, several policy responses (or non-responses) have been motivated using the argumentation that COVID-19 is mostly killing individuals who, even in the absence of COVID-19, would have had only little life years left. However, comprehensive evaluation of the true mortality impact of COVID-19 has not been conducted.

We analyze the premature mortality impact of COVID-19 by calculating the amount of life-years lost across 42 countries covering over 300,000 deaths. We base our analysis on a large recently established and continuously growing database *(4)* and on two different methodological approaches, one based on COVID-19 attributable deaths, and, for selected countries, one based on estimated excess deaths comparing recent mortality levels to an estimated baseline. We are not able to solve the measurement limitations of either of these approaches, but the complementary nature of the two ways of measuring COVID-19 deaths makes these concerns explicit and allows us to evaluate the implications.

## Results

In total, 4,364,326 years of life have been lost to COVID-19 among the studied 42 countries, due to 301,377 deaths from the disease. The average years of life lost per death is 14.5 years. As countries are at different stages of the pandemic trajectory, this study is a snapshot of the impacts of COVID-19 on years of life lost (YLL) as of 13th, June 2020 (for a complete list of countries and their dates at measurement see Supplementary Information). For some countries, such as Italy, transmission rates are low and deaths per day are on a downward trajectory; in such cases, this suggests that the full impacts of the pandemic or at least the first wave of the pandemic are likely captured. However, for other countries still on an upwards incline of transmission rates, the YLL experienced are likely to further increase substantially in the next few months. We encourage context-based interpretation of the results presented here, especially when used for evaluation of the effectiveness of COVID-19 oriented policies.

### Comparisons with other causes of mortality

To put the impacts of COVID-19 on YLL in perspective, we compare it against the premature mortality impacts of three other common causes of death: heart conditions (cardiovascular diseases), traffic accidents (transport injuries), and the seasonal “flu” or influenza (see the Supplementary Materials for the definitions and cause ids). Heart conditions are one of the leading causes of YLL *(5)*, while traffic accidents are a mid level cause of YLL, providing sensible medium and high cause comparison baselines. Finally, common seasonal influenza has been compared against COVID-19, as both are infectious respiratory diseases (though see *(6)*, which suggests vascular aspects to the disease). There is substantial variation in the mortality burden of seasonal influenza by country across years. We compare YLL rates (per 100,000) for COVID-19 against YLL rates for other causes of death; for influenza specifically, YLL rates are for the worst influenza and median influenza years for each country in the period 1990-2017. Comparisons of YLL rates for COVID-19 over YLL rates for other causes are presented in Figure 1.

**Figure 1:**
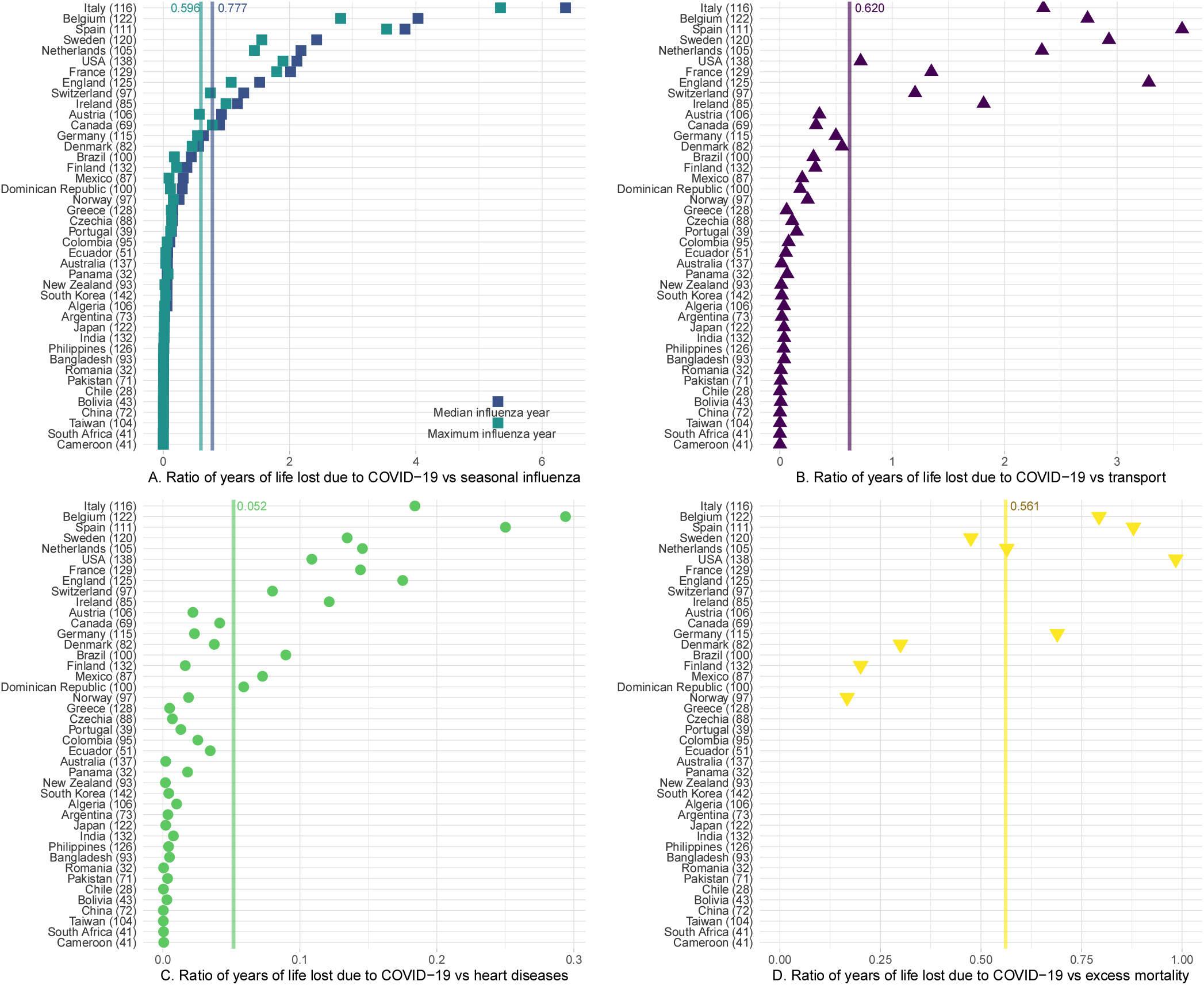
Panels A through C report the ratio of COVID-19 YLL rates over YLL rates from influenza (for median/maximum deadly years for each country), traffic accidents, and heart conditions respectively. Panel D reports, for a subsample of countries with available data, the ratio of YLL rates of COVID-19 deaths over YLL rates of excess deaths. When two causes of mortality affect YLL equally, the ratio is precisely 1; larger ratio values suggest COVID-19 YLL rates are higher than the alternative cause. Average ratios in vertical lines in each panel. See supplementary materials for data and methods.

We find that in heavily impacted highly developed countries, COVID-19 is 2 to 6 times that of the common seasonal influenza, between 1 and 3 times traffic related YLL rates, and between a tenth and a third of the YLL rates attributable to heart conditions. Variation across countries is large, as many countries have YLL rates due to COVID-19 still at very low levels. Results in our Supplementary Materials show that these countries are often countries where relatively few days have passed since first confirmed case of COVID-19.

A noted problem in attributing deaths to COVID-19 has been systematic undercounting of deaths due to COVID-19, as official death counts may reflect limitations in testing as well as difficulties in counting in out-of-hospital contexts. In order to asses the importance of undercounting in our results, we compute excess deaths for 9 countries with available weekly mortality data. A mortality baseline is estimated for each country, sex and age group for weekly all-cause mortality since the first week of 2014. Our results (Figure 1, fourth panel) support the claim that the true mortality burden of COVID-19 is likely to be substantially higher. Comparisons of COVID-19 attributable deaths and excess deaths approaches to calculating YLL suggests that the former on average may underestimate YLL by a factor of 2. Variation across countries is large, and in Belgium, Spain and the United States the two approaches deliver comparable results, but for Denmark, Finland and Norway the excess deaths approach suggests that we may underestimate the YLL by a factor of 3-5.

### Age specific years of life lost

As has been noted early on in the pandemic, mortality rates for COVID-19 are higher for the elderly *(7)*, with postulations that this may be due to correlations with the greater likelihood of these individuals suffering from underlying risk factors *(8)*. This study’s sample presents an average age-at-death of 76 years; yet only a fraction of the YLL can be attributed to the individuals in the oldest age brackets. Globally, 42% of the total YLL can be attributed to the deaths of individuals between 55 and 75 years old, 24% to younger than 55, and 34% to those older than 75. That is, the average figure of 14.5 YLL includes the years lost from individuals close to the end of their expected lives, but the majority of those years are from individuals with significant remaining life expectancy. Across countries, a substantial proportion of YLL can be traced back to the 55-75 age interval, however there remain stark differences in the relative contribution of the oldest and youngest age groups (Figure 2, Panel A). These patterns account for the proportion of YLL for each age group out of the global YLL (see Table S7). In higher income countries, a larger proportion of the YLL is borne by the oldest group compared to the youngest age groups. The opposite pattern appears in low and mid-income countries, where a large fraction of the YLL are from individuals dying at ages 55 or younger.

**Figure 2:**
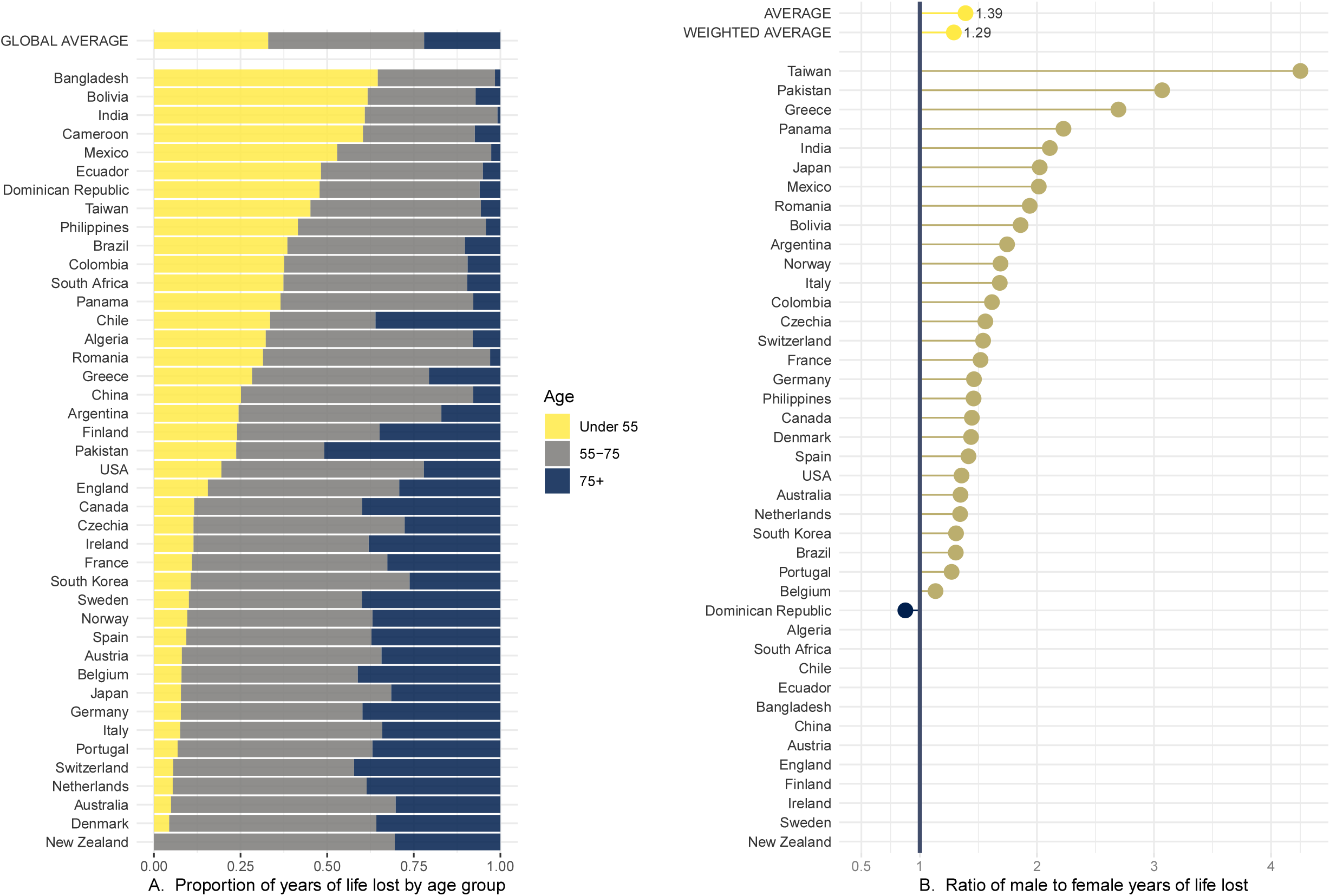
Panel A displays the proportion of YLL that can be traced back to each age group. Panel B reports the ratio of male YLL rates to female YLL rates for the group of countries with available gender specific COVID-19 death counts. Countries with genders equally affected by YLL rate are closer to the parity line at 1, while countries with women more affected have points lying on the left; countries with men more severely affected display points lying to the right. See supplementary materials for data and methods.

### Gender specific years of life lost

It has also become apparent that there are gender disparities in the experience of COVID-19 *(9)*; our study finds this to be true not only in mortality rates, but in absolute years of life lost as well. In the sample of countries for which death counts by gender are available, men have lost 47% more years than women. Two causes directly affect this disparity: 1) a higher average age-at-death of female COVID-19 deaths (73.4 for males, 79 for females), resulting in a relatively lower YLL per death (15.0 and 13.5 for males and females respectively); and 2) more male deaths than female deaths in absolute number (1.3 ratio of male to female deaths).

Though this general pattern is shared by most countries, the size of the disparity varies, as well as the importance of the two above causes. The ratio of male YLL rates (per 100,000) to female YLL rates for COVID-19 spans from near parity, such as in Belgium, to double the YLL rates countries like Mexico (Figure 2, Panel B). For countries that present highly skewed male to female YLL rates (most prevalent in low-income countries), the death count differences across genders contribute the most to this imbalance. Yet, the substantial imbalances remain starkly present among high-income countries as well (see Supplementary Materials for details).

## Discussion

Understanding the full health impact of the COVID-19 pandemic is critical for evaluating the potential policy responses. We analyzed the mortality impact of COVID-19 by calculating the amount of life-years lost across 42 countries covering over 300,000 deaths. From a public health standpoint, years of life lost is crucial in that it assesses how much life has been cut short for populations affected by the disease. We considered COVID-19 attributable deaths throughout in identifying patterns of years of life lost, and as an important robustness check, conducted analysis based on estimated excess deaths comparing recent mortality levels to a (estimated) baseline. Our results deliver three key insights. First, the total years life lost (YLL) as of June 13th, 2020 is 4,364,326, which in heavily affected countries is between 2 to 6 times the median YLL of seasonal influenza or up to a third of heart disease. Second, over two thirds of the YLL are borne by people dying in ages below 75. Third, men have lost 47% more years of life than women.

These results must be understood in the context of an as-of-yet ongoing pandemic and after the implementation of unprecedented policy measures. Existing estimates on the counterfactual of no policy response suggest much higher death tolls and, consequently, YLL. Our calculations based on the projections by *(10)* yield a total impact several orders of magnitude higher, especially considering projections based on a complete absence of interventions (see Supplementary Materials for details on projections). This is in line with further evidence of the life-saving impact of lockdowns and social distancing measures *(11)*.

There are two key sources of potential bias to our results, and these biases operate in different directions. First, COVID-19 deaths may not be accurately recorded, and most of the evidence suggests that on the aggregate level, they may be an undercount of the total death toll. As a result, our YLL estimates may be underestimates as well. We compare our YLL estimates to estimates based on excess death approaches that require more modeling assumptions but are robust to missclassification of deaths. The results of this comparison suggest that on average across countries, we might underestimate COVID-19 YLL rates by a factor of two.

Second, those dying from COVID-19 may be an at-risk population whose remaining life expectancy is shorter than the average person’s remaining life expectancy. This methodological concern is likely to be valid, and consequently our estimate of the total YLL due to COVID-19 may be an overestimate. However, our key results are not the total YLL but YLL ratios and YLL distributions which are relatively robust to the co-morbidity bias. Indeed, this bias also applies to the YLL calculations for the seasonal influenza or heart disease. Thus, the ratio of YLL for COVID-19 compared to other causes of death is more robust to the co-morbidity bias than the estimate on the level of YLL as the biases are present in both the numerator and the denominator. Likewise, the age- and gender distributions of YLL would suffer from serious co-morbidity bias only if these factors vary strongly across the age or gender spectrum.

Some of our findings are consistent with dominant narratives of the COVID-19 impact, others suggest places where more nuanced policy-making can affect how the effects of COVID-19 might be spread among society. Our results confirm that the mortality impact of COVID-19 is large, not only in terms of numbers of death, but also in terms of years of life lost. While the majority of deaths are occurring at ages above 75, justifying policy responses aimed at protecting these vulnerable ages, our results on the age pattern call for heightened awareness of devising policies protecting also the young. The gender differential in years of life lost arises from two components: more men are dying from COVID-19, but men are also dying at younger ages with more potential life years lost than women. Holding the current age distribution of deaths constant, eliminating the gender differential in YLL would require on average a 30% reduction in male death counts; this suggests that gender-specific policies might be equally well justified as those based on age.

## Data Availability

Data and all analyses are available and reproducible one the OSF repository: https://osf.io/5j9nc/?view_only=48f0f69952814e3a8e967370e7b50954

## Acknowledgments

The research funding included grant LCF/PR/GN12/50250002 from La Caixa Foundation for Héctor Pifarré i Arolas and Guillem López Casasnovas. CN is supported by the University of Oxford’s COVID-19 Research Response Fund, number 0009139. The views expressed are those of the author and not necessarily those of the University of Oxford.

## Author contributions

HPA and MM contributed to conceptualization, methodology, supervision, writing original manuscript and review and editing. HPA conducted formal analysis, investigation, data curation. MM contributed data resources. AL conducted formal analysis, data curation, writing original draft, writing review and editing, and visualization. EA and TR conducted data curation and contributed data resources. EA further conducted formal analysis. CN wrote and edited manuscript. GLC contributed to conceptualization, writing and editing of manuscript.

## Data and materials availability

All data and analyses conducted are replicable and available on OSF in the following repository: https://osf.io/5j9nc/?view_only=48f0f69952814e3a8e967370e7b50954.

## Supplementary materials

Materials and Methods

Figs S1 to S11

Tables S1 to S7

References *(12–16)*

